# Identifying real-time surveillance indicators that can be used to estimate COVID-19 hospital admissions

**DOI:** 10.1101/2024.06.25.24309487

**Authors:** Elise N. Grover, Andrew Hill, Irina M. Kasarskis, Emma J. Wu, Nisha Alden, Alicia B. Cronquist, Kirsten Weisbeck, Rachel Herlihy, Elizabeth J. Carlton

## Abstract

Numerous approaches have been used to track COVID-19 trends, from wastewater surveillance to laboratory reporting of diagnostic test results. However, questions remain about how best to focus surveillance efforts during and after public health emergencies. Using an archive of SARS-CoV-2 surveillance data, we reconstructed seven real-time surveillance indicators and assessed their performance as predictors of 7-day moving average COVID-19 hospital admissions in Colorado from October 2020 to March 2024. Models were constructed using neural network models and Ordinary Least Squares regression. We found that hospital census data, emergency-department based syndromic surveillance, and daily COVID-19 hospital admissions were the best indicators of COVID-19 hospital demand in Colorado during the public health emergency (PHE) (October 2020 – May 2023) and after (May 2023 – March 2024). The removal of wastewater from our multi-indicator models resulted in a decrease in model performance, indicating that wastewater provides important and unique information. By contrast, capacity to predict COVID-19 hospital admissions was not meaningfully reduced when sentinel test positivity, statewide test positivity, and/or case report data were dropped from our prediction models during and after the PHE. These findings underscore the importance of hospital-based reporting for monitoring COVID-19 hospital admissions, and, conversely, suggest that case reporting and percent positivity are not essential to monitoring COVID-19 hospitalizations in Colorado.

## Introduction

Ideally, infectious disease surveillance systems provide situational awareness allowing public health and hospital officials to track demand for critical resources during infectious disease outbreaks, and make data-informed decisions in real time^1,2^. For SARS-CoV-2 and other communicable diseases that have the potential to strain healthcare resources, a key interest is developing reliable indicators of COVID-19 hospitalizations. The array of surveillance indicators for SARS-CoV-2 has included healthcare system data (e.g. hospital demand), laboratory-based reporting (percent positivity), case reporting, outbreak reporting, and syndromic and wastewater surveillance. As the public health emergency fades, there are open questions regarding how to allocate limited public health resources to maintain situational awareness for COVID-19 and how best to design surveillance systems for future respiratory disease outbreaks. For example, with changes in disease severity and testing practices, case-based surveillance has become less informative. Many states have ceased case-based surveillance or have de-emphasized it in data analyses while employing combinations of other surveillance methods described above. Furthermore, the Centers for Disease Control and Prevention (CDC) is no longer presenting case-based data on the CDC COVID-19 data website, and as of June 2024, the Council of State and Territorial Epidemiologists has voted to remove COVID-19 from the list of nationally notifiable conditions^3^. As such, evaluating the predictive skill of other surveillance indicators is critically important.

Studies to date have suggested strong positive associations between COVID-19 hospital admissions and SARS-CoV-2 concentrations in wastewater^4–6^, test positivity^7^, and syndromic surveillance^8^. However, a key challenge to evaluating the predictive skill of SARS-CoV-2 surveillance indicators is that many surveillance data sources are reported several days after identification of a reportable condition and data are backfilled as public health officials work to assign cases, hospitalizations and tests to the appropriate date. The use of complete (backfilled) surveillance data in analyses aimed at evaluating the performance of real-time surveillance indicators may over-estimate their predictive skill^9^. In this analysis, we make use of an archive of posted surveillance metrics to re-create what public health officials could see in real-time over the course of the pandemic.

In this study, we assessed the utility of seven different surveillance indicators at predicting COVID-19 hospital admissions between 10/03/2020 and 03/30/2024 in Colorado. Because the end of the public health emergency (PHE) brought shifts in data collection, reporting and testing protocols, we assess surveillance indicator performance for the PHE and post-PHE periods separately, and also evaluated variations in performance over time. The seven surveillance indicators of interest were real-time hospital admissions, hospital census counts, emergency-department-based syndromic surveillance, reported cases, wastewater viral activity level, sentinel test positivity, and statewide test positivity. We assessed the performance of surveillance indicators during the PHE period (10/03/2020 – 05/02/2023) using Long short-term memory (LSTM) neural network models. We then assessed the performance of surveillance indicators during the post-PHE period (05/03/2023 – 03/30/2024) using Ordinary Least Squares (OLS) regression (the shorter time period precluded the use of LSTM models). These analyses are designed to inform how best to allocate limited public health resources to enable situational awareness of COVID-19 hospital demand in the post-PHE period, as well as to inform the design of surveillance systems that promote public health preparedness for future respiratory disease pandemic outbreaks.

## Methods

### Data sources

Our analysis makes use of both real-time and finalized surveillance data. We define finalized data as surveillance data that represents, to the best of our knowledge, the most accurate and complete estimates after data backfill, transfers, and cleaning have taken place. We note that estimates may change over time as data are backfilled and cleaned, particularly later in the time series. We define real-time data are the data visible and retrievable on a given day. The difference between real-time and finalized data may be due to the time it takes to process laboratory and/or surveillance records. For example, real-time case data can include the current estimate of the number of reported cases one day prior, noting that the number of reported cases for that date will eventually be revised over time as reportable surveillance data are cleaned and backfilled. Similarly, on a given day, public health officials may only be able to access SARS-Cov-2 concentrations in wastewater samples collected several days earlier, due to laboratory processing times.

#### Outcome

Daily COVID-19 hospital admissions were our outcome of interest, as this is a key indicator of hospital demand and provides a proxy for incidence^10^. Until 04/30/2024, the federal government required hospitals to report COVID-19 hospital data, herein referred to as HHS Protect^11^. We used finalized hospital admissions data from HHS Protect (05/03/2024 version), as these data are presumed to represent the closest approximation to the true number of patients admitted with COVID-19 on a given day. HHS Protect provides an estimate, for each day, of the number of pediatric and adult patients admitted the previous day with confirmed COVID-19. We used a right-justified 7-day moving average of daily COVID-19 hospital admissions as the outcome variable to smooth random noise and fluctuations in hospital admissions by day of the week.

#### COVID-19 surveillance indicators

We evaluated seven surveillance indicators as potential predictors of COVID-19 hospital admissions (Table 1).

**Table 1.** Description of surveillance measures used in our analysis.

| Indicator | Definition and Source | Data Source | Date Range | Reporting Cadence | Notes |
| --- | --- | --- | --- | --- | --- |
| Hospital Admissions (outcome) | Number of people admitted to a hospital in Colorado the previous day with confirmed COVID-19 (7-day moving average) | HHS Protect | 07/15/2020 – 04/27/2024 | Daily | Finalized data |
| Hospital Admissions (real-time) | Number of people admitted to a hospital in Colorado the previous day with confirmed COVID-19 (7-day moving average) | HHS Protect, posted by CDPHE | 08/20/2020 – 05/02/2023 | Daily until 01/31/2022; M-F until 03/16/2022; Weekly 03/23/2022-05/03/2023* | Real-time data from data publicly posted by CDPHE through the PHE. Real-time data not available post-PHE. |
| Hospital Census | Number of patients currently hospitalized with COVID-19 in a facility in Colorado (daily value) | EMResource (Juvare) provided by CDPHE | 03/22/2020 – 04/16/2024 | Daily through 03/15/2022; Weekly after 03/23/2022* | Real-time data. These data are not backfilled. |
| Syndromic Surveillance | Percent of emergency department visits in the past week with a COVID-19 diagnosis, based on ICD10 codes from participating counties | CDPHE | 01/04/2020 – 05/04/2024 | Weekly | Real-time data. These data are not backfilled. |
| Reported Cases | Number of cases of COVID-19 reported to the state (7-day moving average) | CDPHE – CEDRS | 08/19/2020 – 04/29/2024 | Daily until 02/04/2022; M-F after 02/07/2022 | Real-time data from data publicly posted by CDPHE through the PHE. Real-time data approximated post-PHE using individual-level case reporting records. |
| Wastewater | Wastewater Viral Activity Level, calculated as a weekly flow- and population-normalized concentration using CDC methods | CDPHE | 09/23/2020 – 04/17/2024 | 2 times per week | Finalized data. Suppressed by four days to account for laboratory and data processing times. |
| Sentinel Test Positivity | Percentage of NAAT and antigen tests at 29 participating facilities that return a positive result over a 7-day period | CDPHE | 09/27/2020 – 04/28/2024 | Weekly | Real-time data. These data are not backfilled. |
| Statewide Test Positivity | Percentage of NAAT tests from all facilities and laboratories in Colorado, including CDPHE, that return a positive result (7-day moving average) | ELR | 08/19/2020 – 05/08/2024 | Daily through 02/11/2022; M-F 02/14/2022 through 05/09/2023* | Real-time data from data publicly posted by CDPHE through the PHE. Real-time data approximated post-PHE using individual-level laboratory reporting records. |

\*When COVID hospital admission data and statewide positivity reporting shifted to weekdays and then once a week, these files always included daily values. In contrast, when hospital census reporting shifted to weekly, only one estimate per week was available.
\*\*WVAL is calculated on a per-site basis, with each site usually reporting 2x per week. The per-site values are averaged to create a single weekly per-site value. These per-site weekly values are then averaged again across all sites to generate a single state-wide weekly WVAL metric.

To reconstruct real-time case reports, hospital admissions (PHE only), and statewide test positivity (PHE only), we retrieved data from an archive of publicly posted files from the Colorado Department of Public Health & Environment (CDPHE) (available: https://drive.google.com/drive/folders/1efQVBclGxwnCCYVLbzH96A0QRSWwmTYI). For case reports during the PHE, datasets were posted daily, and each dataset contained the number of cases of COVID-19 reported to the state from 03/05/2020 through the day before the file was posted. We then used these datasets to reconstruct the number of cases that had been reported 1 to 14 days prior to a given date, as known on that date. A similar approach was used to reconstruct real-time hospital admissions data during the PHE, extracting data from 1 to 14 days prior. Statewide positivity was posted daily, and then Monday through Friday, and included 7-day moving average positivity. These files were used to extract the 7-day moving average positivity posted on a given day, representing the real-time estimate of current statewide positivity. This archive ended on 05/02/2023, with the end of the PHE.

For case report data post-PHE, we retrospectively estimated real-time case counts using the Colorado Electronic Disease Reporting System (CEDRS), which includes a timestamp indicating both the case report date and the date the case record was added to the surveillance system. Using these timestamps, we calculated the cumulative sum of cases for each report date that were known for each calendar date in order to approximate real-time data. We used a similar approach to estimate real-time statewide positivity, using individual-level statewide positivity records that included a timestamp for the date the record was created. Real-time hospital admissions could not be reconstructed post-PHE and were therefore excluded from post-PHE models. We smoothed both COVID-19 hospital admissions and reported cases by applying a right-justified 7-day moving average.

COVID-19 sentinel test positivity and COVID-19 hospital census data were not backfilled and thus represent real-time data in their final state. In March 2022, the hospital census data reporting requirements changed from daily reporting to reporting on Tuesdays each week. Syndromic surveillance may experience minor backfill over time, primarily due to changes in the denominator, but these changes rarely meaningfully change estimates; as such, we consider the final versions of these data reasonable approximations of real-time data.

SARS-CoV-2 concentrations in wastewater were posted twice weekly by the Colorado Wastewater Surveillance Program. Values were posted for each participating facility, with dates reflecting sample collection dates. We used this information to estimate a normalized, weekly statewide concentration, using per-facility population and flow-rate information provided by CDPHE and the CDC’s Wastewater Viral Activity Level (WVAL) algorithm (available: https://www.cdc.gov/nwss/about-data.html), retrieved in April 2024. We note that only finalized wastewater data were available To approximate real-time data for any given day, we hid the most recent four days of data which accounts for the lab processing time between sample collection, laboratory processing and data posting.

For all indicators, missing data were filled with the last observed value to reflect likely real-time data management practices. All analyses were conducted for the period 10/03/2020 – 03/30/2024 unless otherwise indicated. Data were cleaved on 03/30/2024 to ensure stability of recently reported data for analysis. We defined the PHE period as 10/03/2020 – 05/02/2023 (while the PHE ended 05/11/2023, changes to the format for publicly posted data began several days earlier, which is why we end the period on 05/03/2023). We define the post PHE period as 05/03/2023 – 03/30/2024.

Further details on the data sources and construction process used for COVID-19 hospital admissions and each of the surveillance indicators included in our analyses are included in the Supplementary Information.

The Colorado Multiple Institution Review Board reviewed this study and determined it to be exempt from Institutional Review Board review.

### Statistical Analysis

We conducted two preliminary analyses. First, we estimated the lags between each surveillance indicator and 7-day average COVID-19 hospital admissions using cross-correlation methods^7,12^. Each surveillance measure captures a different state in the disease process and is subject to different reporting efficiencies; as such, some surveillance measures are leading indicators of COVID-19 hospital admissions and others are synchronous or lagging indicators. For each surveillance indicator, we evaluated lags from 13 days before to 13 days after COVID-19 hospital admission date. Each indicator was log-transformed and the cross-correlation calculated. We selected the lag with the observed maximum correlation value between the two time series; if the observed maximum correlation value for a surveillance indicator was observed at multiple timepoints, we selected the lag closest to zero. A positive value (t + x days) indicates a lagging indicator that follows hospital admissions, while a negative value (t – x days) represents a leading indicator of hospital admissions by “x” days. Cross-correlations were calculated once for the PHE period and once for the post-PHE period. Cross-correlations could not take into account daily changes in data backfill. Therefore, we used finalized data for surveillance indicators that were subject to backfill (reported cases, wastewater, statewide positivity post PHE) and real-time data when estimates were not changed over time (hospital census, syndromic surveillance, sentinel test positivity, statewide positivity during the PHE). We excluded real-time hospital admissions from this analysis because these data are subject to backfill and finalized hospital admissions is the outcome of interest.

Second, we visualized how the relationships between each of the seven surveillance indicators and COVID-19 hospital admissions varied over time. We used the *NonParRolCor* package in R^13^ to estimate pairwise Spearman correlation coefficients between each surveillance indicator and 7-day moving average COVID-19 hospital admissions over a 90-day rolling window throughout the course of the pandemic. The mean correlation (ρ) was estimated for the PHE and post-PHE periods for each indicator vs. the outcome. To account for the lags between surveillance indicators and COVID-19 hospital admissions, we used the results of the lags analysis for the full time period: syndromic surveillance (t – 1 day); 7-day moving average of daily case reports (t – 3 days); wastewater viral activity levels (t – 3 days); weekly sentinel test positivity (t – 7 days); and 7-day moving average of statewide test positivity (t – 4 days) (Supplementary Table S1). Hospital census was found to be a lagging indicator; however, as we were interested in information known on or before a given date, we applied a zero-day lag. We also applied a zero-day lag to real-time COVID-19 hospital admissions, which could not be included in the aforementioned lags analysis.

#### Surveillance indicator performance during the PHE

To assess the predictive skill of the COVID-19 surveillance indicators during the PHE, we evaluated the performance of a range of single and multiple predictor models using artificial neural network (ANN) models. ANN is a machine learning computational model inspired by the biological neural networks of the human brain, wherein nodes communicate via connections similar to the way that neuron cells communicate via axons and dendrites^14,15^. During the ANN model training process, the weight of each node connection is adjusted to minimize the difference between the actual and predicted output via the network^15^. In this analysis, we employed Long Short-Term Memory (LSTM) models^16^, a type of ANN model specifically designed for handling time-series prediction tasks with long term temporal dependencies.

In order to train and test our models, we divided our data into training and test sets, fitting the model using the training data and evaluating the model using the testing data. The data used in each of our LSTM prediction models were broken into three equal length series of data folds: Fold 1 (10/03/2020 – 08/08/2021), Fold 2 (08/09/2021 – 06/20/2022), and Fold 3 (06/21/2022 – 05/02/2023). Each time fold contained one smaller and one larger peak in COVID-19 hospitalization admissions (Figure 1), though the magnitude and length of these peaks varied between the three folds. We employed a three-fold cross validation process wherein each model was run three times using three different training/testing splits: Split 1) Training on Fold 2 & 3 and Testing on Fold 1; Split 2) Training on Folds 1 & 3 and Testing on Fold 2; and Split 3) Training on Folds 1 & 2 and Testing on Fold 3.

**Figure 1.**
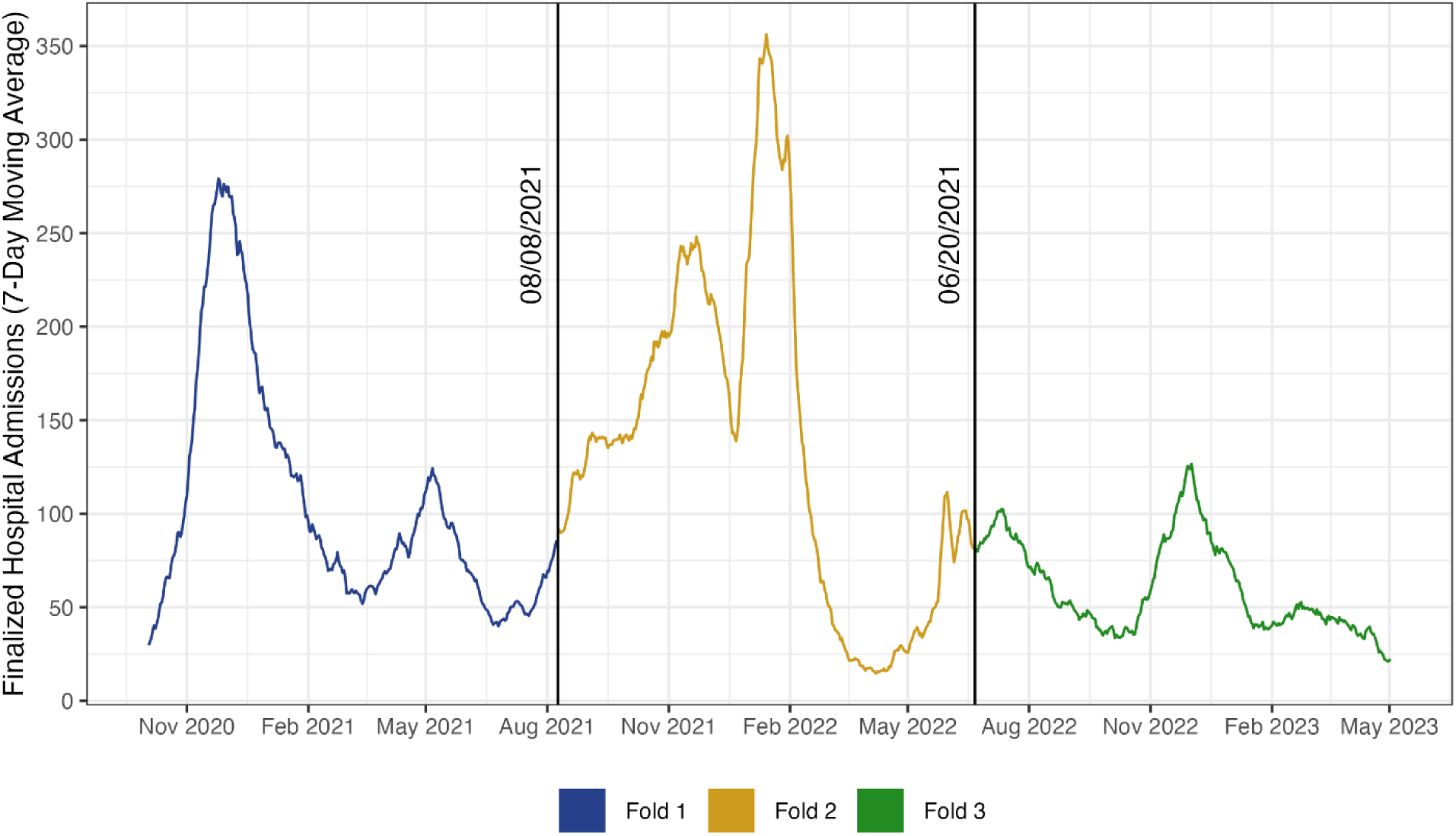
COVID-19 hospital admissions over time and the three divisions used for a 3-fold cross validation of the neural network models used in the analyses of the public health emergency period. Each time fold contains one smaller and one larger peak in COVID-19 hospitalization admissions. Fold 1 spans the time period from 10/03/2020 – 08/08/2021; Fold 2 spans the period of 08/09/2021 – 06/20/2022; and Fold 3 spans the time period from 06/21/2022 – 05/02/2023.

Lags between surveillance indicators and COVID-19 hospital admissions were defined using the observed maximum correlation with finalized hospital admissions identified in our cross-correlation analyses. We used a zero-day lag when the analysis indicated it was a lagging indicator and assigned a lag of t – 1 days for real-time hospital admissions. We note that LSTM models have the flexibility to identify alternate, longer lags as part of the learning process. The seven COVID-19 surveillance indicators assessed in our LSTM models were as follows: 1) 7-day moving average daily hospital admissions (t – 1 days); 2) daily hospital census (t – 0 days); 3) weekly syndromic surveillance (t – 2 days); 4) 7-day moving average case reports (t – 1 days); 5) biweekly wastewater viral activity level (t – 3 days); 6) weekly sentinel test positivity (t – 6 days); and 7) 7-day moving average statewide test positivity (t – 4 days).

Each surveillance indicator was initially assessed independently on their ability to predict COVID-19 hospital admissions across the three training-testing splits. The Mean Absolute Error (MAE) was calculated for each test fold, estimating the average number of hospital admissions that our predicted values differed from the actual value on a given day was estimated across each test fold. We then estimated the average error across all three test folds as the mean MAE. Each surveillance indicator was then ranked from 1^st^ (best) to 7^th^ (worst) on its ability to independently predict daily hospital admissions across the PHE period.

We then constructed a full model, using all seven indicators, and evaluated the degree to which our model performance would suffer in their ability to accurately predict 7-day average COVID-19 hospital admissions when a surveillance indicator was removed from the model. As above, we estimated the mean MAE for each model. We then estimated the change in model error for each leave-one-out model relative to the full models:

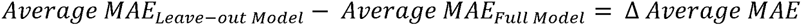

A positive Δ average MAE indicates that, on average, the leave-one-out model performed worse than the full model (and ostensibly, the omitted indicator is improving the predictive performance of the model), whereas a negative Δ average MAE indicates that the model performed better without the omitted indicator. As above, we ranked the models from best (1^st^) to worst. This analysis also includes one model in which both case reports and statewide positivity were removed as the reporting requirements for these indicators might change in the fall of 2024. We also ran a model excluding both sentinel and statewide positivity, to assess the general performance of laboratory reporting data.

#### Surveillance indicator performance after the PHE

The limited number of days in the post-PHE period (05/03/2023 – 03/30/2024) meant that splitting the data into training and testing datasets would have left LSTM models underpowered. As an alternative, we developed and compared a range of OLS regression models to assess the predictive skill of COVID-19 surveillance indicators during the post-PHE period. During this period, six COVID-19 surveillance indicators were available and assessed on their ability to accurately and reliably predict 7-day moving average COVID-19 hospital admissions (real-time hospital admissions were not available). As above, we used the results of the cross-correlation analysis to define lags between indicators and COVID-19 hospital admissions. The six surveillance indicators assessed were as follows: hospital census (t – 0 days); syndromic surveillance (t – 1 days); reported cases (t – 0 days); wastewater viral activity level (t – 0 days); sentinel test positivity (t – 13 days); and statewide test positivity (t – 13 days).

As above, we initially assessed the ability of each surveillance indicator to independently predict COVID-19 hospital admissions, and then evaluated performance in full, leave-one-out, and leave out cases and statewide positivity models. To compare the goodness of fit between models containing each individual surveillance metric, we recorded each model’s adjusted R^2^, AIC, and BIC estimates. The model (i.e., surveillance indicator) with the best goodness of fit, as indicated by the highest adjusted R^2^, was ranked first, while the model with the lowest R^2^ was ranked last. In the event of a tie, we used the AIC/BIC to rank the models (where a lower AIC/BIC resulted in a higher rank).

## Results

Figure 2 shows the distribution of finalized 7-day average COVID-19 hospitalizations and the seven surveillance indicators.

**Figure 2.**
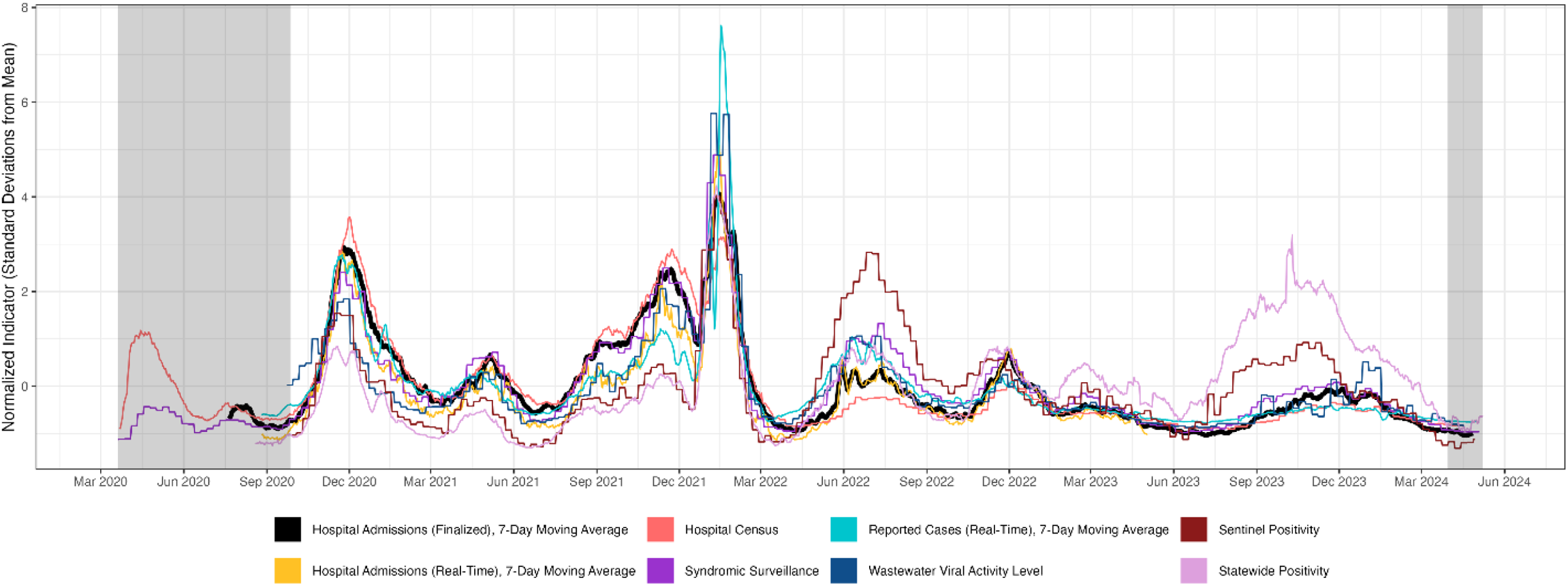
Finalized 7-day moving average COVID-19 hospital admissions (thick line) and seven real-time COVID-19 surveillance indicators (colorful lines), Colorado, 03/20/2020 – 05/08/2024. The prediction analyses performed for this study used only time periods where data for indicators were simultaneously available. Grey shaded regions indicate time periods excluded from the analysis of surveillance predictors: from 03/20/2020 – 09/27/2020 not all surveillance indicators were available; 03/30/2024 – 05/08/2024 represents a period during which all or most surveillance indicators were available, but data were cleaved at 03/30/2024 to ensure stability of recently reported data for analysis. For the purpose of visualization, all indicators have been z-score normalized and reported as number of standard deviations from the mean.

### Lags between surveillance indicators and COVID-19 hospital admissions

Our analysis estimated that all surveillance indicators led COVID-19 hospital admissions in the PHE and post-PHE period except hospital census (both periods), cases (post-PHE), and wastewater viral activity level (post-PHE). (Table 2, Supplementary Figures S1 – S4). The correlation coefficients between COVID-19 hospital admissions and reported cases, syndromic surveillance and wastewater viral activity levels were generally high (> 0.7) in the two weeks preceding hospital admissions, suggesting that these indicators lead hospital admissions. Notably, the estimated lead times were shorter in the post-PHE period, and the estimated lags for both wastewater viral activity levels and reported cases were zero days. In contrast, the estimated lags for sentinel and statewide positivity were much longer in the post-PHE period (13 days), compared to 6 (sentinel) and 4 (statewide) days during the PHE, and the correlations were weaker (range: 0.50 – 0.70) during the PHE. Hospital census was not a leading indicator of hospital admissions; during the PHE, the estimated lag was zero days, and in the post-PHE period, the estimated lag was seven days.

**Table 2.** Estimated lags between six surveillance indicators and 7-day average COVID-19 hospital admissions, during the PHE and post-PHE periods.

| Surveillance indicator | Estimated lag (days), PHE<br>(07/21/2020* – 05/02/2023) | Estimated lag (days),<br>post-PHE (05/03/2023 –<br>03/30/2024) |
| --- | --- | --- |
| Case reports (finalized), 7-day moving average | -3 | 0 |
| Hospital census (real-time) | 0 | 7 |
| Wastewater Viral Activity Level (finalized) | -3 | 0 |
| Syndromic surveillance (real-time) | -2 | -1 |
| Sentinel test positivity (real-time) | -6 | -13 |
| Statewide test positivity (real-time PHE, finalized post-PHE), 7-day moving average | -4 | -13 |

PHE = public health emergency
Lags were estimated using cross-correlation analysis from 13 days before to 13 days after COVID-19 hospital admission date. The lag was defined using the maximum correlation. In the case of a tie, values closest to zero were used. A positive lag corresponds to indicators that lagged admissions (time + x days), negative lags correspond to indicators that lead admissions (time - x days). Lags with a value of 0 indicate that the correlation between 7-day average hospital admissions and the surveillance indicator was strongest on the same day (i.e., no lag).
\*Start date was later for the following indicators: wastewater viral activity level (09/23/2020), sentinel test positivity (09/27/2020), and statewide test positivity (08/19/2020).

### Correlations over time

Hospital-based indicators, including real-time COVID-19 hospital admissions, hospital census and emergency-room based sentinel surveillance were most closely correlated with COVID-19 hospital admissions during and after the PHE (Figure 3, Supplementary Table S2). Reported cases and wastewater viral activity levels were also strongly correlated with COVID-19 hospital admissions (mean correlation during the PHE 0.78 and 0.76, respectively). Sentinel and statewide positivity were less well correlated with COVID-19 hospital admissions. The rolling correlation between each surveillance indicator and COVID-19 hospital admissions was generally weaker during the post-PHE period. The rolling correlation dipped for many indicators during the first Omicron wave (beginning late 2023), but this was not observed for wastewater viral activity level.

**Figure 3.**
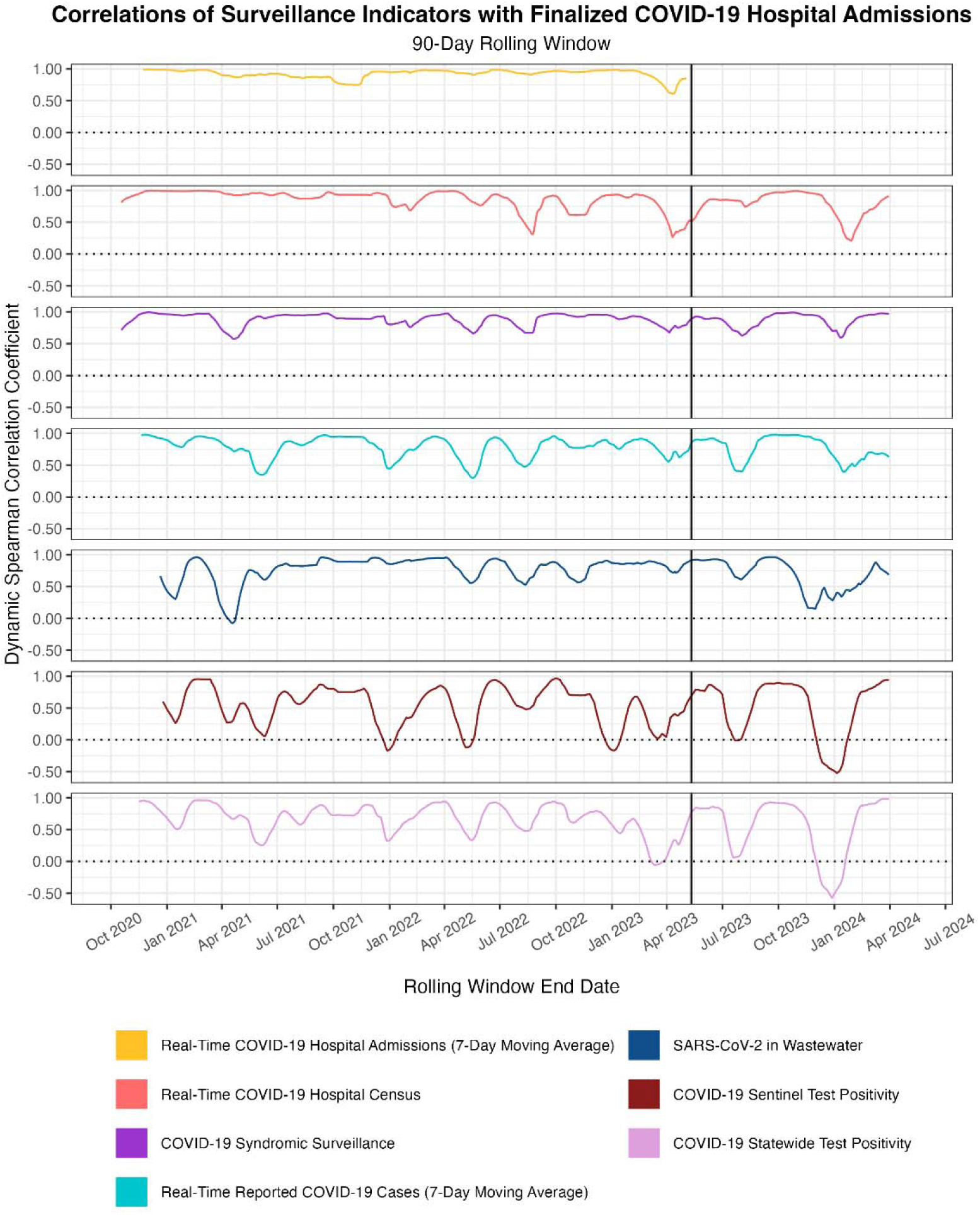
Dynamic Spearman correlations between 7-day moving average finalized COVID-19 hospital admissions and seven surveillance indicators over a 90-day right-justified rolling window through 03/30/2024. Start dates vary by indicator, based on first availability. To account for differential timing between certain surveillance indicators and finalized hospital admissions, we shifted leading surveillance indicators according to lead times determined by a cross-correlation analysis. Lagging indicators were not shifted. For visualization, each pair of indicators was scaled to the same y-axis by applying a z-score normalization to transform the data to the number of standard deviations from the mean. Horizontal dotted lines represent a correlation coefficient of zero. Vertical solid lines represent the end of the COVID-19 public health emergency (PHE) on 05/11/2023. *Date ranges and lags applied for each surveillance indicator are as follows: real-time COVID-19 hospital admissions (7-day moving average), to 05/03/2023 with no lag applied; real-time COVID-19 hospital census, 07/21/2020 to 03/30/2024 with no lag applied; COVID-19 syndromic surveillance, 07/21/2020 to 03/30/2024 with a 1-day lead applied; real-time reported COVID-19 cases, 08/23/2020 to 03/30/2024 with a 3-day lead applied; SARS-CoV-2 in wastewater, 09/23/2020 to 03/30/2024 with a 3-day lead applied; COVID-19 sentinel test positivity, 09/27/2020 to 03/30/2024 with a 7-day lead applied; and COVID-19 statewide test positivity, 08/19/2020 to 03/30/2024 with a 4-day lead applied.

### Surveillance indicator model performance during the PHE

The independent predictor LSTM models, designed to assess the ability of 7 surveillance indicators to independently predict finalized 7-day moving average daily hospital admissions during the PHE, indicated that real-time hospital census (t – 0 days), real-time hospital admissions (t – 1 days), and real-time syndromic surveillance (t – 2 days) were the 3 best performing predictors, ranked 1^st^, 2^nd^, and 3^rd^ respectively (Table 3). Individual surveillance indicators tended to have higher model accuracy when predicting hospitalizations in Test Fold 1 (10/03/2020 – 08/08/2021) and Test Fold 3 (06/21/2022 – 05/02/2023), and the lowest model accuracy (i.e., highest MAE) tended to arise when predicting hospitalizations for Test Fold 2 (08/09/2021 – 06/20/2022). One exception to this finding was for our worst performing surveillance indicators (ranked 6^th^ and 7^th^), weekly sentinel positivity and 7-day moving average statewide positivity. In both cases, the lowest model accuracy was observed when we used information from Folds 1 & 2 to train models to predict daily hospital admissions during Fold 3.

**Table 3.** The performance of seven surveillance indicators in predicting 7-day moving average COVID-19 hospital admissions during the COVID-19 public health emergency, from 10/03/2020 – 05/02/2023.

| Model description | Test Fold 1 Scaled MAE | Test Fold 2 Scaled MAE | Test Fold 3 Scaled MAE | Average MAE <sup>a</sup> | Model Rank by Average MAE <sup>b</sup> | $\Delta$ Average MAE <sup>c</sup> |
| --- | --- | --- | --- | --- | --- | --- |
| **Individual surveillance indicator models:** |  |  |  |  |  |  |
| **Name of included indicator** |  |  |  |  |  |  |
| Real-time hospital admissions (t – 1 days) | 13.650 | 31.900 | 27.318 | 24.289 | 2 | NA |
| Real-time hospital census (t – 0 days) | 14.245 | 24.542 | 14.597 | 17.795 | 1 | NA |
| Real-time syndromic surveillance (t – 2 days) | 28.334 | 28.078 | 21.361 | 25.924 | 3 | NA |
| Real-time case reports (t – 3 days) | 14.066 | 55.061 | 21.807 | 30.311 | 4 | NA |
| Wastewater viral activity level (t – 3 days) | 33.440 | 78.501 | 43.664 | 51.868 | 5 | NA |
| Real-time sentinel positivity (t – 6 days) | 31.695 | 68.015 | 104.948 | 68.219 | 6 | NA |
| Real-time statewide positivity (t – 4 days) | 42.505 | 60.876 | 117.026 | 73.469 | 7 | NA |
| **Leave-indicator(s)-out models:** |  |  |  |  |  |  |
| **Name of excluded indicator** |  |  |  |  |  |  |
| Full model (Include all indicators) | 18.901 | 24.835 | 8.746 | 17.494 | 5 | 0 |
| Real-time hospital admissions (t – 1 days) | 12.119 | 33.796 | 10.907 | 18.941 | 8 | 1.447 |
| Real-time hospital census (t – 0 days) | 22.048 | 36.490 | 12.895 | 23.811 | 10 | 6.317 |
| Real-time syndromic surveillance (t – 2 days) | 15.891 | 28.902 | 9.508 | 18.100 | 6 | 0.606 |
| Real-time case reports (t – 3 days) | 21.764 | 26.414 | 7.414 | 18.531 | 7 | 1.037 |
| Wastewater viral activity level (t – 3 days) | 17.284 | 28.699 | 11.352 | 19.112 | 9 | 1.618 |
| Real-time sentinel positivity (t – 6 days) | 10.187 | 18.367 | 6.029 | 11.528 | 1 | -5.966 |
| Real-time statewide positivity (t – 4 days) | 11.780 | 26.218 | 8.056 | 15.351 | 3 | -2.143 |
| Real-time case reports (t – 3 days) & statewide positivity (t – 4 days) | 10.551 | 24.795 | 10.114 | 15.153 | 2 | -2.341 |
| Real-time sentinel positivity (t – 6 days) & real-time statewide positivity (t – 4 days) | 14.076 | 24.870 | 10.812 | 16.586 | 4 | -0.908 |

MAE = Mean Absolute Error
NA = Not applicable. The $\Delta$ in Average MAE was only calculated for the leave-indicator(s)-out models
<sup>a</sup> Average of the MAE across the three models
<sup>b</sup> Each model's average MAE across the three-time folds was used to rank model performance from best (1<sup>st</sup>), indicated by the lowest three-fold average MAE, to worst, indicated by the highest three-fold average MAE. Rankings were performed separately for the individual surveillance indicator models and the leave-indicator(s)-out models.
<sup>c</sup> This is estimated as the difference between each leave-indicator-out model average MAE, and the full model's average MAE (leave-one-out model average MAE - full model average MAE). The higher the average $\Delta$ MAE, the more influential the left-out predictor. Models that have a positive $\Delta$ MAE indicate that the model without the left-out predictor had an overall increase in error and therefore performed worse without the omitted predictor, whereas a negative $\Delta$ MAE indicate that the model performed better without the omitted predictor.

In the leave-one out models, we observed the greatest loss in predictive performance relative to the full model in those that excluded daily hospital census data (Δ Average MAE = +6.3), wastewater viral activity level (Δ Average MAE = +1.6) and real-time hospital admissions (Δ Average MAE = +1.4). Removal of syndromic surveillance and case reports led to modest declines in model performance. Notably, several models performed better than the full model when an indicator was removed: this was true for models that excluded sentinel positivity, statewide positivity, both sentinel and statewide positivity, and both statewide positivity and case reports. These results indicate that neither sentinel positivity, statewide positivity, nor the combination of statewide positivity and case reports add to the predictive performance of the full model.

### Surveillance indicator model performance after the PHE

The independent predictor OLS models, designed to assess the ability of 6 surveillance indicators to independently predict finalized 7-day moving average daily hospital admissions after the PHE, indicated that real-time syndromic surveillance was the top performing model (R^2^ = 0.933), followed by real-time hospital census (R^2^ = 0.907) and real-time case reports (R^2^ = 0.897) (Table 4). By contrast, we found lower goodness of fit for the models using statewide positivity (R^2^ = 0.697), sentinel positivity (R^2^ = 0.629) and wastewater (R^2^ = 0.592), which were ranked 4^th^, 5^th^ and 6^th^, respectively.

**Table 4.**
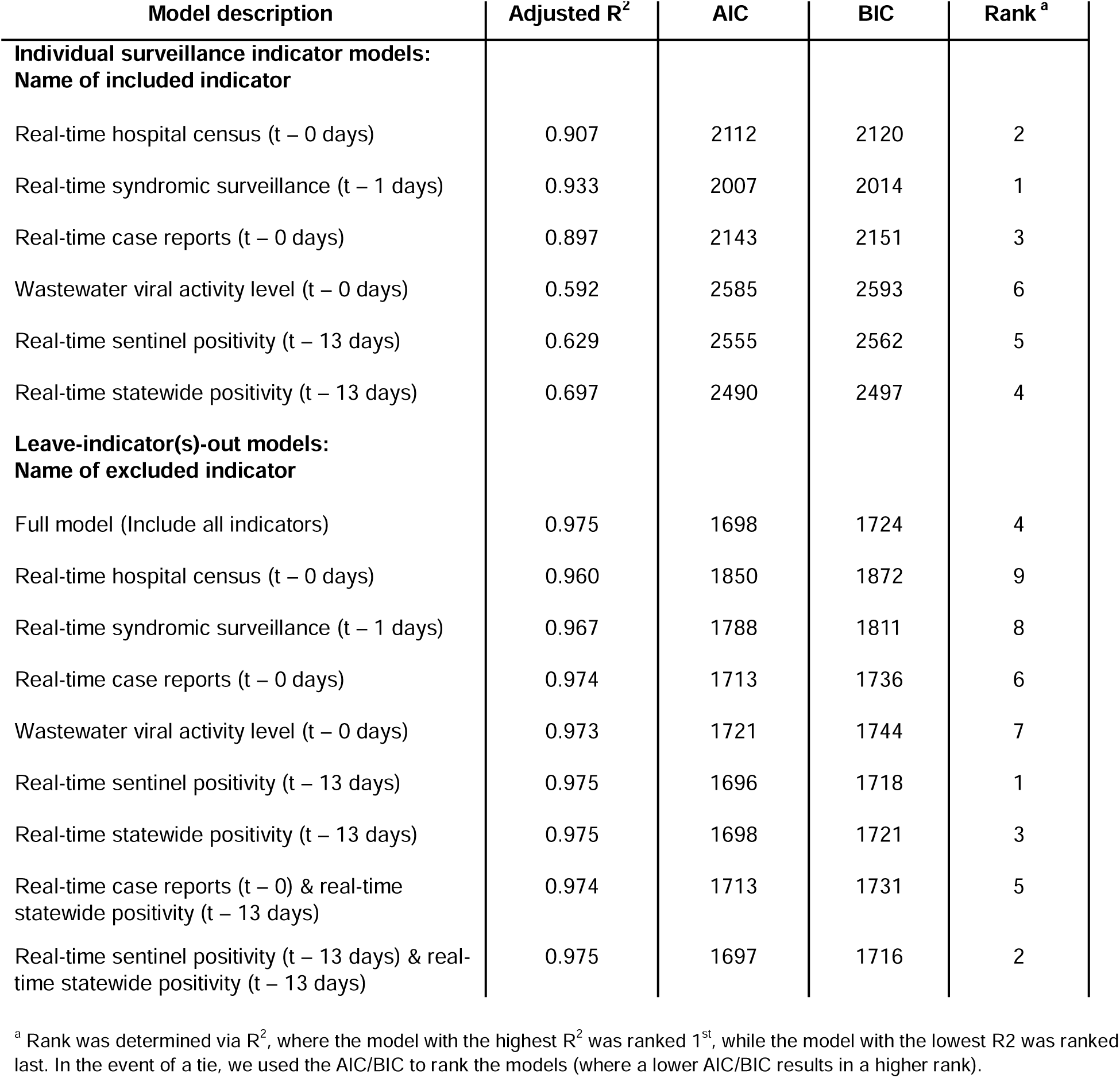
The performance of six real-time surveillance indicators in predicting 7-day moving average COVID-19 hospital admissions after the COVID-19 public health emergency, from 05/03/2023 – 03/30/2024.

| Model description | Adjusted R<sup>2</sup> | AIC | BIC | Rank<sup>a</sup> |
| --- | --- | --- | --- | --- |
| **Individual surveillance indicator models:** |  |  |  |  |
| **Name of included indicator** |  |  |  |  |
| Real-time hospital census (t – 0 days) | 0.907 | 2112 | 2120 | 2 |
| Real-time syndromic surveillance (t – 1 days) | 0.933 | 2007 | 2014 | 1 |
| Real-time case reports (t – 0 days) | 0.897 | 2143 | 2151 | 3 |
| Wastewater viral activity level (t – 0 days) | 0.592 | 2585 | 2593 | 6 |
| Real-time sentinel positivity (t – 13 days) | 0.629 | 2555 | 2562 | 5 |
| Real-time statewide positivity (t – 13 days) | 0.697 | 2490 | 2497 | 4 |
| **Leave-indicator(s)-out models:** |  |  |  |  |
| **Name of excluded indicator** |  |  |  |  |
| Full model (Include all indicators) | 0.975 | 1698 | 1724 | 4 |
| Real-time hospital census (t – 0 days) | 0.960 | 1850 | 1872 | 9 |
| Real-time syndromic surveillance (t – 1 days) | 0.967 | 1788 | 1811 | 8 |
| Real-time case reports (t – 0 days) | 0.974 | 1713 | 1736 | 6 |
| Wastewater viral activity level (t – 0 days) | 0.973 | 1721 | 1744 | 7 |
| Real-time sentinel positivity (t – 13 days) | 0.975 | 1696 | 1718 | 1 |
| Real-time statewide positivity (t – 13 days) | 0.975 | 1698 | 1721 | 3 |
| Real-time case reports (t – 0) & real-time statewide positivity (t – 13 days) | 0.974 | 1713 | 1731 | 5 |
| Real-time sentinel positivity (t – 13 days) & real-time statewide positivity (t – 13 days) | 0.975 | 1697 | 1716 | 2 |

<sup>a</sup> Rank was determined via R<sup>2</sup>, where the model with the highest R<sup>2</sup> was ranked 1<sup>st</sup>, while the model with the lowest R<sup>2</sup> was ranked last. In the event of a tie, we used the AIC/BIC to rank the models (where a lower AIC/BIC results in a higher rank).

We found high goodness of fit comparing the full model (all six post-PHE indicators) to each of the leave-out-indicator(s) models (R^2^ >0.950 in all models). The three worst-performing models were those that left out hospital census data (R^2^ = 0.960), syndromic surveillance data (R^2^ = 0.967), and wastewater data (R^2^ = 0.973), respectively. The adjusted R^2^ of the full model and the models that excluded sentinel positivity, statewide positivity, and both sentinel and statewide positivity were the same (R^2^ = 0.975), though a comparison of model AIC and BIC indicated slightly better performance in the leave-out sentinel positivity model (AIC = 1696, BIC = 1718), the leave-out both sentinel and statewide positivity model (AIC = 1697, BIC = 1716), and the leave-out statewide positivity model (AIC = 1698, BIC = 1721), as compared to the full model (AIC = 1698, BIC = 1724). Models that excluded reported cases and models that excluded both reported cases and statewide positivity were essentially equivalent to the full model (R^2^ = 0.974 vs. 0.975).

## Discussion

Our findings suggest that hospital-based indicators, such as emergency-department based syndromic surveillance and hospital census counts, are some of the best predictors of actual 7-day moving average COVID-19 hospital admissions. The single-predictor models that included these indicators were among the best performing single-predictor models both during and after the PHE. Additionally, removing these indicators from multi-predictor models resulted in a notable loss in model performance, evidence that these measures are important predictors in composite models. Not surprisingly, real-time hospital admissions was among the top ranked surveillance indicators during the PHE (this metric could not be reconstructed during the post-PHE period), which is, logically, a close approximation to actual (finalized) COVID-19 hospital admissions. In contrast, we found that reported cases, sentinel test positivity, and statewide test positivity were consistently lower-performing surveillance indicators in both the individual indicator models and the composite models for the PHE and post-PHE periods.

We undertook this analysis because we were interested in identifying which indicators are critical to estimating COVID-19 hospital demand. This is particularly relevant now, as the US federal government and states and territorial public health agencies reevaluate how to monitor COVID-19 trends and severity^10^. We found that our capacity to predict COVID-19 hospital admissions does not change substantially when both statewide test positivity and case report data are excluded from our prediction models, nor when statewide and/or sentinel positivity are excluded. We caution that the focus of our analyses was statewide COVID-19 hospital admissions; indicators such as case reports and laboratory-based reporting may provide situational awareness on aspects such as testing demand, and high-burden populations beyond the scope of this analysis.

Others have found that wastewater viral concentrations can be used to forecast COVID-19 hospital admissions and hospital demand at state and county levels^6,17,18^. We found that wastewater was among the lower performing surveillance indicators in individual predictor models (ranked 5^th^ during the PHE, 6^th^ in the post-PHE period), but the removal of wastewater from multi-indicator models resulted in a notable decrease in model performance, indicating that removing wastewater results in the loss of some important and/or unique information. We note that our findings are consistent with prior work that found wastewater outperformed test positivity in predicting hospital admissions^5^.

Key strengths of our analysis include the ability to reconstruct real-time surveillance data using publicly posted daily records, the ability to examine the performance of multiple surveillance indicators alone and in combination, and the long time series over which we conducted this analysis (October 2020 to March 2024). Reporting requirements, clinical practice and test-seeking behavior changed markedly over the past four years. For example, in Colorado, hospitals tested all patients for SARS-CoV-2 until March 2022, at which point patients admitted with but not for COVID-19 were less likely to be captured in hospital census and admissions counts. Likewise, increasing availability and use of home testing kits and a general decrease in COVID-19 illness severity over time were also likely sources of decreased case ascertainment across the study period. We were therefore surprised by the consistency of our findings between the PHE and post-PHE periods. Generally, the PHE models performed worse during the second period, which included both the most virulent variant (Delta) and the variant that caused the greatest number of infections (Omicron BA.1), highlighting the challenges of surveillance when there are large shifts in virus characteristics.

There were a few noteworthy limitations in this analysis. First, this analysis was designed to evaluate the performance of COVID-19 surveillance indicators beginning in October 2020, nearly seven months after the World Health Organization declared a global pandemic. The data requirements to provide situational awareness in the early weeks of an emerging infectious disease event are beyond the scope of this analysis. In Colorado, case reports were among the few data available in March of 2020, and were critical to providing situational awareness and supporting modeling efforts until hospital admission data became consistently available^19^. Second, the post-PHE timeframe was significantly shorter than the PHE period, meaning we had insufficient data to employ neural network models to evaluate surveillance indicator performance. Nonetheless, our post-PHE OLS models had high goodness-of-fit (e.g., R^2^ > 0.8) for several of the individual surveillance indicator models, as well as for all of our multi-predictor models. Third, while we did our best to recreate data that public health officials could see in real time, we were unable to recreate real-time hospital admissions in the post PHE period. We relied on approximations for reported cases and statewide positivity post-PHE (we did this using individual-level data with reporting timestamps), and for wastewater during both the PHE and post-PHE periods (we did this by blinding data for four days to account for laboratory processing times). Nevertheless, we consider the use of real-time surveillance indicators or their approximations a novel contribution of this analysis, that has not been well covered in the literature.

## Conclusion

Hospital-based surveillance indicators, including daily hospital census and emergency-department based syndromic surveillance, as well as daily reporting of COVID-19 hospital admissions are some of the strongest indicators of COVID-19 hospital demand in Colorado currently, and during much of the PHE. Our analysis suggests the discontinuation of SARS-CoV-2 case reporting and statewide laboratory surveillance should not negatively impact the ability to estimate COVID-19 hospital demand in the post-PHE era. As public health agencies refine surveillance strategies for monitoring COVID-19, they should consider integrating these findings into their overall plans.

## Acknowledgements

The authors would like to thank Drs. Jude Bayham, Talia Quandelacy and Jonathan Samet for providing input on the design and interpretation of the analyses

## Data availability

All data used in these analyses was obtained from third parties or public resources. Request for access to these data can be directed to the following parties: the Colorado Department of Public Health & Environment (syndromic surveillance, hospital census, case reports, wastewater and sentinel positivity data); and HHS Protect (hospital admissions data).

## Author contributions

All authors contributed the formulation of the research design. NA, ABC, RH and KW provided the data used in these analyses and advised on data curation. AH, IMK and EJW performed the data curation, cleaning, and statistical analyses. EG, EJC prepared the first draft of the manuscript. All authors contributed to the interpretation of results and contributed substantial edits to the draft. All authors have read and agreed to the published version of the manuscript.

## References

1 Stoto, M. A., Rothwell, C., Lichtveld, M. & Wynia, M. K. A National Framework to Improve Mortality, Morbidity, and Disparities Data for COVID-19 and Other Large-Scale Disasters. American journal of public health 111, S93–S100 (2021). 10.2105/AJPH.2021.306334

2 Lipsitch, M. & Santillana, M. Enhancing Situational Awareness to Prevent Infectious Disease Outbreaks from Becoming Catastrophic. Curr Top Microbiol Immunol 424, 59–74 (2019). 10.1007/82_2019_172

3 CDC. National Notifiable Diseases Surveillance System, <https://www.cdc.gov/nndss/index.html> (2024).

4 Hopkins, L. et al. Citywide wastewater SARS-CoV-2 levels strongly correlated with multiple disease surveillance indicators and outcomes over three COVID-19 waves. The Science of the total environment 855, 158967 (2023). 10.1016/j.scitotenv.2022.158967

5 Hill, D. T. et al. Wastewater surveillance provides 10-days forecasting of COVID-19 hospitalizations superior to cases and test positivity: A prediction study. Infect Dis Model 8, 1138–1150 (2023). 10.1016/j.idm.2023.10.004

6 Galani, A. et al. SARS-CoV-2 wastewater surveillance data can predict hospitalizations and ICU admissions. The Science of the total environment 804, 150151 (2022). 10.1016/j.scitotenv.2021.150151

7 Scobie, H. M. et al. Correlations and Timeliness of COVID-19 Surveillance Data Sources and Indicators - United States, October 1, 2020-March 22, 2023. MMWR Morb Mortal Wkly Rep 72, 529–535 (2023). 10.15585/mmwr.mm7219e2

8 Levy, M. J. et al. Correlation between Emergency Medical Services Suspected COVID-19 Patients and Daily Hospitalizations. Prehosp Emerg Care 25, 785–789 (2021). 10.1080/10903127.2020.1864074

9 Reich, N. G. et al. A collaborative multiyear, multimodel assessment of seasonal influenza forecasting in the United States. Proc Natl Acad Sci U S A 116, 3146–3154 (2019). 10.1073/pnas.1812594116

10 Silk, B. J. et al. COVID-19 Surveillance After Expiration of the Public Health Emergency Declaration - United States, May 11, 2023. MMWR Morb Mortal Wkly Rep 72, 523–528 (2023). 10.15585/mmwr.mm7219e1

11 Centers for Disease Control and Prevention (CDC). COVID-19 Hospital Data Reporting, <https://www.cdc.gov/nhsn/covid19/hospital-reporting.html> (2024).

12 Reich, N. G. et al. Assessing the utility of COVID-19 case reports as a leading indicator for hospitalization forecasting in the United States. medRxiv (2023). 10.1101/2023.03.08.23286582

13 Polanco-Martínez, J. M. & López-Martínez, J. L. NonParRolCor: An R package for estimating rolling correlation for two regular time series. Softwarex 22 (2023). ARTN 101353 10.1016/j.softx.2023.101353

14 James, G., Witten, D., Hastie, T. & Tibshirani, R. An introduction to statistical learning. Vol. 112 (Springer, 2013).

15 Choi, R. Y., Coyner, A. S., Kalpathy-Cramer, J., Chiang, M. F. & Campbell, J. P. Introduction to Machine Learning, Neural Networks, and Deep Learning. Transl Vis Sci Technol 9, 14 (2020). 10.1167/tvst.9.2.14

16 Zhang, J., Zhu, Y., Zhang, X., Ye, M. & Yang, J. Developing a Long Short-Term Memory (LSTM) based model for predicting water table depth in agricultural areas. Journal of Hydrology 561, 918–929 (2018). 10.1016/j.jhydrol.2018.04.065

17 Li, X. et al. Wastewater-based epidemiology predicts COVID-19-induced weekly new hospital admissions in over 150 USA counties. Nature communications 14, 4548 (2023). 10.1038/s41467-023-40305-x

18 Schenk, H. et al. Prediction of hospitalisations based on wastewater-based SARS-CoV-2 epidemiology. The Science of the total environment 873, 162149 (2023). 10.1016/j.scitotenv.2023.162149

19 Buchwald, A. G. et al. Estimating the Impact of Statewide Policies to Reduce Spread of Severe Acute Respiratory Syndrome Coronavirus 2 in Real Time, Colorado, USA. Emerg Infect Dis 27, 2312–2322 (2021). 10.3201/eid2709.204167

