## Supplementary Information for "Identifying real-time surveillance indicators that can be used to estimate COVID-19 hospital admissions"

**Supplementary Methods: Detailed information on surveillance indicators.**

***Real-Time COVID-19 Hospital Admissions.*** Real-time COVID-19 hospital admissions represent the non-revised number of pediatric and adult patients admitted the previous day with confirmed COVID-19. Data were sourced from HHS Protect (<https://www.cdc.gov/nhsn/covid19/hospital-reporting.html>) and posted publicly by CDPHE from 08/20/2020 – 05/03/2023. Datasets were posted daily from 08/20/2020 – 01/31/2022; Monday through Friday from 02/01/2022 – 03/16/2022; and weekly from 03/23/2022 – 05/03/2023. Weekly files still included hospital admissions data for each calendar day. We used these datasets to reconstruct the number of COVID-19 hospital admissions reported 1 to 14 days prior to the date the dataset was published. When datasets transitioned to longer reporting intervals, missing days were filled with the last observed value. We smoothed real-time hospital admission data by applying a right-justified 7-day moving average. Because only finalized COVID-19 hospital admissions data is available after the public health emergency, real-time hospital admissions data were not included in analyses after the end of the public health emergency.

***COVID-19 Hospital Census.*** COVID-19 hospital census represents the number of patients currently hospitalized with confirmed COVID-19 at a facility in Colorado on a given day. Data was obtained from EMResource (Juvare) and publicly posted by CDPHE from 03/22/2020 – 04/16/2024. Data were posted daily from 03/22/2020 – 03/15/2022; and weekly from 03/22/2022 forward. After 3/22/2022, hospital census estimates were only submitted for Tuesdays; other days in the week did not have a value. Hospital census data reported by hospitals through EMResource were posted daily by CDPHE in real-time, but data are not backfilled, so the value reported on a given date did not change over time. To achieve a daily timestep, missing days were filled with the last observed value.

***Syndromic Surveillance.*** COVID-19 syndromic surveillance reports the percentage of emergency department (ED) visits in the past week with a COVID-19 discharge diagnosis, with ICD10 codes (U07.1 and J12.82). The Colorado Syndromic Surveillance Program includes facilities in 14 counties: Adams, Arapahoe, Boulder, Broomfield, Denver, Douglas, El Paso, Jefferson, La Plata, Larimer, Mesa, Montezuma, Pueblo, and Weld Counties. Data were reported weekly at the end of each MMWR week from 01/04/2020 to present day. For analysis, we included data through 03/30/2024. Data are subject to lags or spikes in values due to delays in health facility reporting, and data may be incomplete for the most recent week; however, data are not backfilled. To achieve a daily timestep, missing days were filled with the last observed value until the next posted value.

***Reported Cases.*** Reported cases represent the number of confirmed cases of SARS-CoV-2 by date reported via the Colorado Electronic Disease Reporting System (CEDRS) at CDPHE. We used publicly posted files of daily reported case counts posted from 08/20/2020 – 05/09/2023. Cases were reported daily from 08/20/2020 – 07/16/2021; Monday through Friday from 07/19/2021 – 08/13/2021, daily from 08/16/2021 – 02/04/2022; Monday through Friday from 02/07/2022 – 05/09/2023. All posted files included the number of reported cases for each calendar date. There were data gaps during these periods. Data were missing for the periods from 08/13/2022 – 08/29/2022, and from 10/28/2022 – 11/12/2022. These were filled using a dataset containing daily individual-level reported cases, received from CDPHE in April 2024. To achieve daily estimates from this data source we calculated the cumulative sum for each report date that would be available on a specific calendar day using timestamps in the dataset for report date and case creation date.

Archived real-time reported state-wide case data were not available after 05/09/2023, so we retrospectively created daily reported case data from a line list of all reported cases from CEDRS obtained from CDPHE on 04/30/2024. This dataset was used to generate estimates of real-time reported cases for the time period 05/10/2023 – 03/30/2024, using the same method for the missing data in 2022.

***SARS-CoV-2 in Wastewater.*** Wastewater data were obtained from the Colorado Wastewater Surveillance Program. Data are posted publicly twice a week by utility (available: <https://cdphe.maps.arcgis.com/apps/dashboards/d79cf93c3938470ca4bcc4823328946b>). To generate an estimate of state-wide wastewater trends, we used the above public dataset in conjunction with per-facility population and flow-rate information provided by CDPHE to generate flow-population normalized time series for each facility. For each facility and measurement, flow normalized measurements were calculated by multiplying the raw concentration by the flow rate, and then dividing by the population served by that facility.  After normalization, we applied a second normalization step, based on the CDC’s Wastewater Viral Activity Level (WVAL) algorithm (available: <https://www.cdc.gov/nwss/about-data.html>). This normalization transforms concentration values into standard deviations from a baseline calculated from the 10^th^ percentile of historical wastewater measurements from the previous 12 months. This step reduces issues with comparing/aggregating values between facilities and between reporting stages. After normalization, we created weekly values for each facility/reporting stage by taking the median of the recorded values and generate statewide weekly values by taking the median across the weekly values from all facilities. After normalization, these values are averaged over time to generate a state-wide aggregate time series following methods provided by CDC. For analysis, observations were lagged four days to account for the time between sample collection and report date. To achieve a daily timestep, missing days were filled with the last observed value.

***COVID-19 Sentinel Test Positivity.*** COVID-19 sentinel test positivity represents the percentage of COVID-19 diagnostic tests given at sentinel sites over a 7-day period that return a positive result. The Colorado COVID-19 sentinel surveillance testing system includes participating facilities across the state that administer both nucleic acid amplification tests (NAAT) and antigen tests. NAAT is defined as molecular assays, primarily reverse transcription polymerase chain reaction (RT-PCR). Sentinel test positivity data is reported weekly at the end of each MMWR week. Sentinel test positivity data were obtained from CDPHE and include data from 09/27/2020 – 04/28/2024. Sentinel positivity was calculated for the entire state as the sum of positive NAAT and antigen tests divided by the sum of total NAAT and antigen tests across all participating facilities for each MMWR week. We included missing or unnamed facilities that still had a valid count. If either total or positive test count was missing from a facility during a given week, that facility’s data was excluded for that week. Test positivity for the weeks of 11/29/2020 and 12/27/2020 were missing from the dataset; for these weeks, we used the value from the prior week. Data are not backfilled. To achieve a daily timestep, missing days were filled with the last observed value.

***COVID-19 Statewide Test Positivity.*** COVID-19 statewide test positivity is the percentage of NAAT COVID-19 diagnostic tests performed that return a positive result, calculated as a 7-day moving average. Rapid antigen tests are not reported in this dataset. Data were obtained from CDPHE through ELR and include laboratory reports from CDPHE’s state laboratory and commercial laboratories in Colorado. During the PHE, laboratories were required to report all NAAT results, including positive, negative, and inconclusive results,^[1]^ and at the time of writing, positive and negative NAAT results in most settings in the state of Colorado are still required to be reported to CDPHE.^[2]^ For statewide positivity before the end of the PHE, we used archived daily files, posted publicly on a daily basis from 08/20/2020 – 02/11/2022 and Monday through Friday from 02/14/2022 – 05/09/2023. Although datasets were not posted daily after 02/11/2022, each archived dataset still included daily test positivity values as well as 7-day moving average values. Archived datasets were combined and the first reported value for 7-day moving average positivity for each date was retained to represent what was known in real time. To extend the timeline beyond 05/09/2023, we supplemented real-time statewide positivity data with an individual-level statewide positivity dataset obtained via a one-time data transfer from CDPHE. This dataset spans 03/08/2020 – 05/08/2024 and comprises approximately 23 million observations, each observation intended to represent a unique SARS-CoV-2 NAAT testing event. Antigen tests are not included in this dataset. To ensure that each individual appeared a maximum of once on a given day, we removed replicate observations with identical names, dates of birth, collection dates, result dates, and create dates. Collection date was defined as the date on which the sample was collected; result date is defined as the date on which the result became available; create date is defined as the date on which the result was entered into the ELR database. Aggregating by create date, as opposed to collection or result date, ensured that data reflected what was known in real time. Statewide test positivity was calculated as the sum of all positive COVID-19 tests by create date divided by the sum of all COVID-19 tests by create date. We smoothed all statewide positivity data after the end of the PHE by applying a right-justified weighted 7-day moving average.

**Supplementary Table S1.** Estimated lags between six surveillance indicators and 7-day average COVID-19 hospital admissions, across the full PHE and post-PHE periods, 07/21/2020 – 03/30/2024. The start date was later for the following indicators: wastewater viral activity level (09/23/2020), sentinel test positivity (09/27/2020), and statewide test positivity (08/19/2020).

| **Surveillance indicator** | **Estimated lag (days), full study period*** |
| --- | --- |
| Cases (finalized), 7-day moving average | -3 |
| Hospital census, daily | 1 |
| Wastewater Viral Activity Level | -3 |
| Syndromic Surveillance | -1 |
| Sentinel Testing | -7 |
| Statewide Positivity | -4 |

Lags were estimated using cross correlation analysis from 13 days prior to 13 days post COVID-19 hospital admission date. The lag was defined using the maximum correlation. In the case of a tie, values closest to zero were used. A positive lag corresponds to indicators that lagged admissions (time + x days), negative lags correspond to indicators that lead admissions (time – x days). Lags with a value of 0 indicate that the correlation between 7-day average hospital admissions and the surveillance indicator was strongest on the same day (i.e., no lag).

**Supplementary Table S2.** Mean correlations between surveillance indicators and finalized hospital admissions over a 90-day rolling window, during and after PHE. Start dates vary by indicator, based on first availability. For the purpose of analysis, we define the PHE period as ending 05/02/2023 and post-PHE period ran from 05/03/2023 – 03/30/2024.

| **Time Series 1** | **Time Series 2** | **Mean correlation (PHE)*** | **Mean correlation (Post-PHE)*** |
| --- | --- | --- | --- |
| Finalized hospital admissions* | Real-time hospital admissions* | 0.920 | N/A |
|  | Real-time hospital census | 0.861 | 0.778 |
|  | Syndromic surveillance (t – 1) | 0.882 | 0.867 |
|  | Real-time reported cases* (t – 3) | 0.783 | 0.764 |
|  | Wastewater (t – 4) | 0.771 | 0.684 |
|  | Sentinel positivity (t – 7) | 0.525 | 0.501 |
|  | Statewide positivity* (t – 4) | 0.667 | 0.547 |

*given as 7-day moving average

**Supplementary Figure S1.** Cross-correlations between reported cases and 7-day moving average hospital admissions up to 13 days prior to and after hospital admissions. Both daily case counts and 7-day moving average cases are shown. Darker colors indicate the full time period, medium colors indicate the public health emergency (PHE) period, and lighter colors indicate the post PHE. The height of the lines indicates the correlation between the two time-series by different lags on the x-axis. The maximum cross-correlations for each surveillance indicator are marked with a circle.

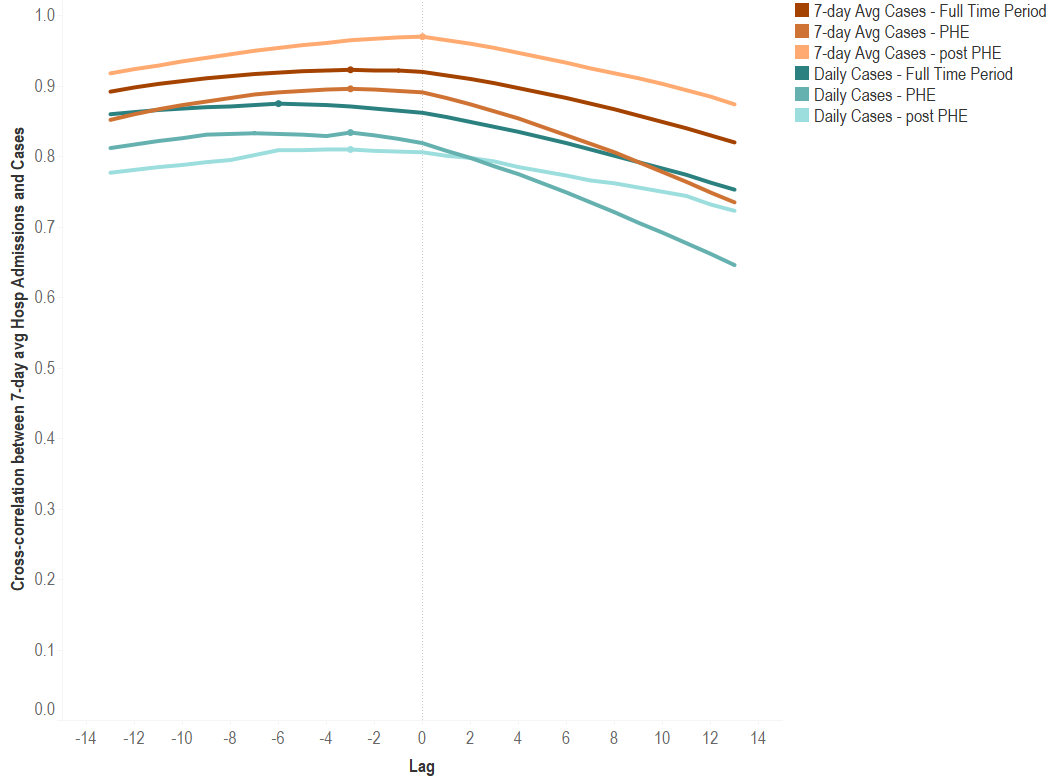

**Supplementary Figure S2.** Cross-correlations between daily count of COVID-19 hospitalized patients (hospital census) and 7-day moving average hospital admissions up to 13 days prior to and after hospital admissions. Both daily sense and 7-day average COVID-19 hospital census counts are shown. Darker colors indicate the full time period and lighter colors indicate the public health emergency. The height of the lines indicates the correlation between the two time-series by different lags on the x-axis. The maximum cross-correlations for each surveillance indicator are marked with a circle.

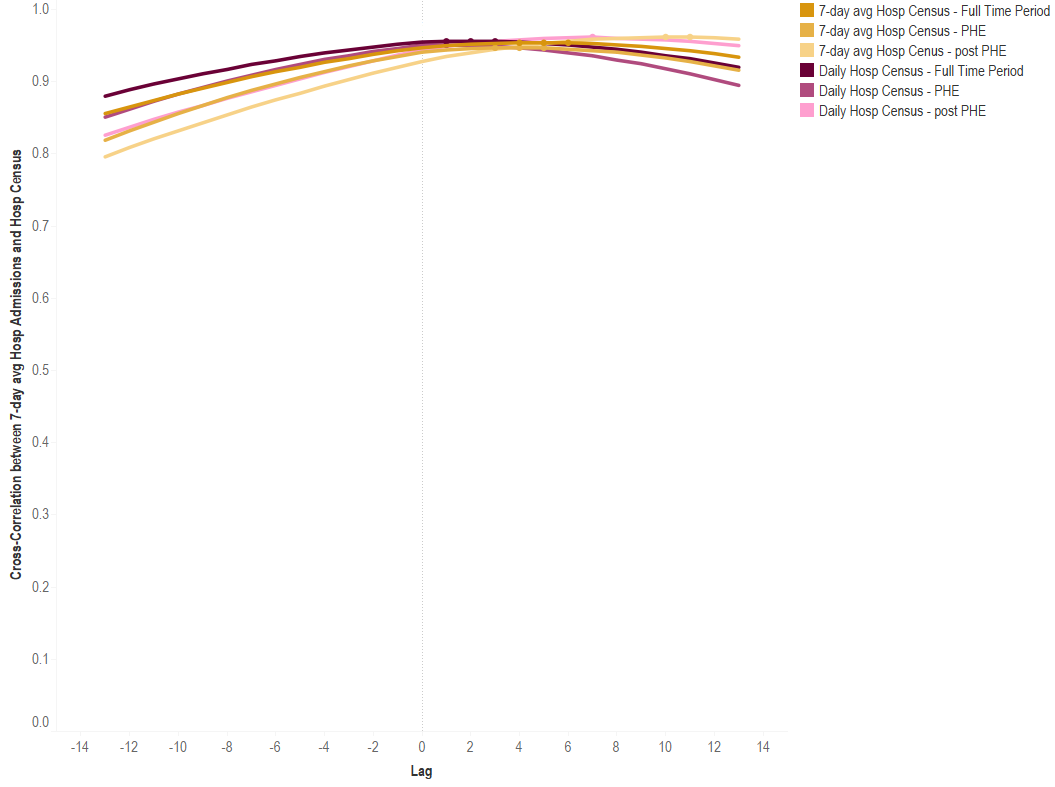

**Supplementary Figure S3.** Cross-correlation between testing indicators and 7-day moving average hospital admissions up to 13 days prior to and after hospital admissions. Both 7-day moving average state-wide test positivity and 7-day average sentinel test positivity are shown. Darker colors indicate the full time period and lighter colors indicate the public health emergency. The height of the lines indicates the correlation between the two time-series by different lags on the x-axis. The maximum cross-correlations for each surveillance indicator are marked with a circle.

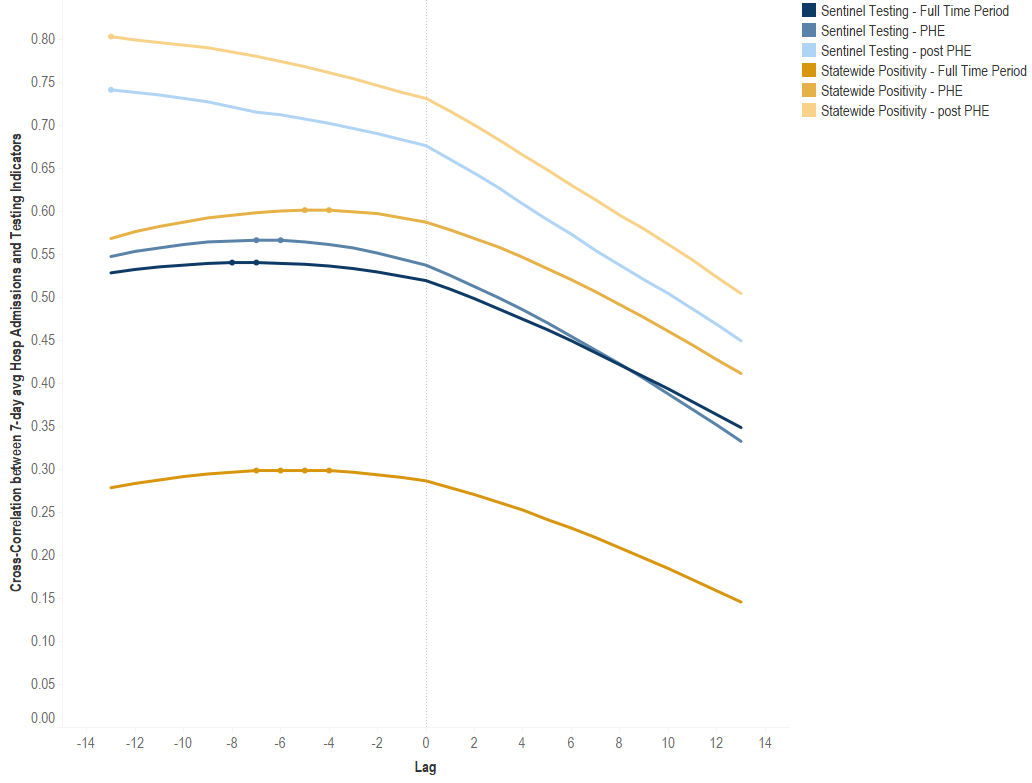

**Supplementary Figure S4.** Cross-correlation between weekly wastewater viral activity levels, weekly syndromic surveillance and 7-day moving average hospital admissions up to 13 days prior to and after hospital admissions. Darker colors indicate the full time period and lighter colors indicate the public health emergency. The height of the lines indicates the correlation between the two time-series by different lags on the x-axis. The maximum cross-correlations for each surveillance indicator are marked with a circle.

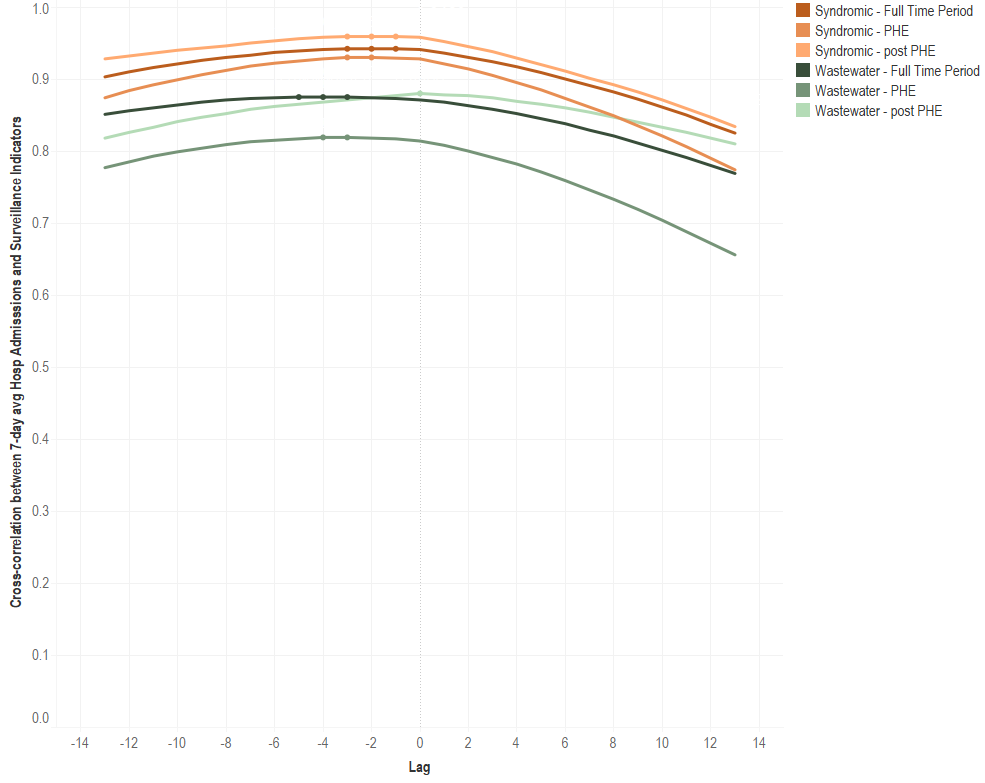

1. CARES Act (Public Law 116-136, § 18115(a),) (2022).
2. CDPHE. COVID-19 (SARS-CoV-2) Reporting Requirements. Available: <https://cdphe.colorado.gov/report-a-disease/covid> 2024.
